# Trends and prediction in daily incidence and deaths of COVID-19 in the United States: a search-interest based model

**DOI:** 10.1101/2020.04.15.20064485

**Authors:** Xiaoling Yuan, Jie Xu, Sabiha Hussain, He Wang, Nan Gao, Lanjing Zhang

**Author notes:** Correspondence: Lanjing Zhang, MD, Department of Pathology, Princeton Medical Center, 1 Plainsboro Rd., Plainsboro, NJ 08563. Drs Yuan and Xu made equal contributions to the works, and should be considered as co-first authors. Conflict of Interest Disclosures: No disclosures were reported.

## Abstract

**Background and Objectives:** The coronavirus disease 2019 (COVID-19) infected more than 586,000 patients in the U.S. However, its daily incidence and deaths in the U.S. are poorly understood. Internet search interest was found correlated with COVID-19 daily incidence in China, but not yet applied to the U.S. Therefore, we examined the association of internet search-interest with COVID-19 daily incidence and deaths in the U.S.

**Methods:** We extracted the COVDI-19 daily incidence and death data in the U.S. from two population-based datasets. The search interest of COVID-19 related terms was obtained using Google Trends. Pearson correlation test and general linear model were used to examine correlations and predict future trends, respectively.

**Results:** There were 555,245 new cases and 22,019 deaths of COVID-19 reported in the U.S. from March 1 to April 12, 2020. The search interest of COVID, “COVID pneumonia,” and “COVID heart” were correlated with COVDI-19 daily incidence with ∼12-day of delay (Pearson’s r=0.978, 0.978 and 0.979, respectively) and deaths with 19-day of delay (Pearson’s r=0.963, 0.958 and 0.970, respectively). The COVID-19 daily incidence and deaths appeared to both peak on April 10. The 4-day follow-up with prospectively collected data showed moderate to good accuracies for predicting new cases (Pearson’s r=-0.641 to −0.833) and poor to good accuracies for daily new deaths (Pearson’s r=0.365 to 0.935).

**Conclusions:** Search terms related to COVID-19 are highly correlated with the trends in COVID-19 daily incidence and deaths in the U.S. The prediction-models based on the search interest trend reached moderate to good accuracies.

---

**C**oronavirus disease 2019 (COVID-19) has been pandemic in the world.^1-4^ It has infected more than 560,000 of Americans.^3,5^ Several attempts were successfully made to model COVID-19 daily incidence in China.^1,6^ However, the trends of daily incidence and deaths of COVID-19 in the U.S. are still poorly understood. Recently, internet search interest was found correlated with daily incidence of COVID-19 in China, with the lagging time of 8 to 10 days.^7^ Google search interest was also used to track or model COVID-19 trends in Europe, Iran and Taiwan.^8-10^ Indeed, internet search interest has been used for modelling and detecting influenza epidemics in the U.S. and Australia.^11,12^ We therefore aimed to examine the association of search interest with daily incidence and death of COVID-19 in the U.S., using population-based data and a semi-parametric model.

## Methods

The daily incidence and new deaths of COVID-19 in the U.S. were extracted from the 1-point-3-acres.com^5^ and the Johns Hopkins COVID-19 data repository^3^ on April 9, 2020, respectively, for modelling. We later obtained additional data from these sites to evaluate our model’s accuracy using Pearson’s correlation coefficients. Data from the World Health Organization situation-reports appeared significantly inconsistent, and thus were not used.^13^ According to the 1-point-3-acres.com website, their data were extracted from various media and government websites, and have been manually verified,^5^ and have been used by various parties including Johns Hopkins COVID-19 data repository, World Health Organization, and many others. Due to the use of publicly available, de-identified data and lack of protected health information, the study is exempt from an Institutional Board Review (Category 4).

We used the Google trends function to extract the data of search interest with search period of March 1 to April 10, 2020 and COVID-19 related search terms. The terms we used were COVID-19, COVID, coronavirus, pneumonia, “High temperature,” cough, “Covid heart,” “Covid pneumonia” and “Covid diabetes.” Google trends search interest represented search interest relative to the highest search interest for the given time and region.^7,12^ A value of 100 is the peak popularity for the term, while a score of 0 means there were not enough data for this term.

We then examined the lag correlations of the terms’ search interests with COVID-19 daily incidence and new deaths as described before.^7^ The date-shifts of our interest were up to 20 for daily incidence and 23 for daily death, respectively. The terms with the top-3 correlation coefficients were used to build respective generalized linear models. Based on these models, we used the existing search interests to predict future COVID-19 daily incidence and new deaths in the U.S., which would be compared with the prospectively collected data for assessing prediction accuracies.

All statistical analyses will be carried out using Stata (version 15). The models’ accuracies were assessed using Pearson’s r. All P values were 2-sided. Only a P<0.05 was considered statistically significant.

## Results

The Johns Hopkins data repository and 1-point-3-acres.com provided slightly different estimates of COVID-19 daily incidence and deaths in the U.S., although they shared data. The data of given dates from 1-point-3-acres.com dataset varied by the release dates. Considering the data inconsistency, we chose the John Hopkins data for modelling. The 1-point-3-acres.com data were used in a sensitivity study. There were 555,245 new cases and 22,019 deaths of COVID-19 reported in the U.S. from March 1 to April 12, 2020, with a crude mortality of 3.97%.

Google Trends search interests had a 2-day delay in reporting (i.e. a search on April 8 yielded data up to April 6). COVID-19 has a much lower search interest score than COVID (**Figure 1**), and was excluded from additional analysis owing to its close relationship with COVID. As reported before, the correlation coefficients of search terms changed with lagging time (**Figure 2**). Among the 9 terms we searched, COVID, “COVID pneumonia” and “Covid heart” had the top-3 correlation coefficients for correlation with daily incidence and new deaths (**Table**). Our predicted COVID-19 daily incidence and new cases would plateau for about 12 days (**Figure 3**), suggesting a possible 12-day plateau of these epidemiologic parameters in the future. The sensitivity study using 1-point-3-acres data revealed the correlation coefficients that were similar to those produced using Johns Hopkins data (**Table**). The 4-day followup with prospectively collected data show moderate to good accuracies for predicting new cases (Pearson’s r ranged −0.641 to −0.833) and poor to good accuracies for new deaths (Pearson’s r ranged 0.365 to 0.935).

**Table.**
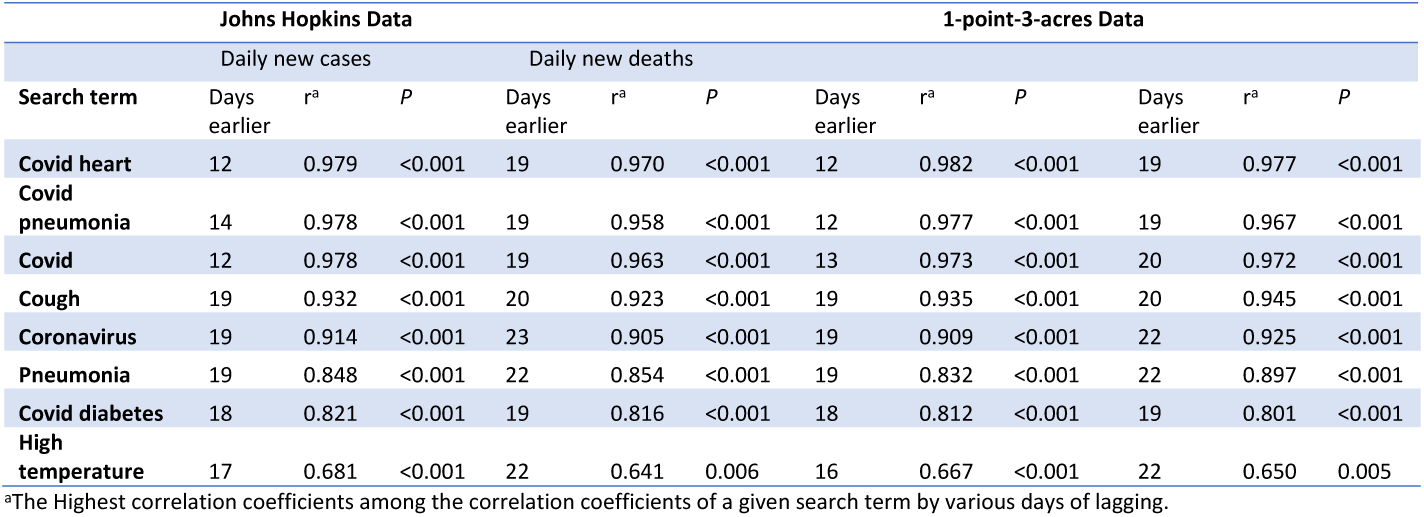
The search term of the top-3 correlation coefficients for correlations with COVID-19 daily incidence and deaths, March 1 to April 8, 2020

| Search term | Johns Hopkins Data |  |  |  |  |  | 1-point-3-acres Data |  |  |  |  |  |
| --- | --- | --- | --- | --- | --- | --- | --- | --- | --- | --- | --- | --- |
|  | Daily new cases |  |  | Daily new deaths |  |  |  |  |  |  |  |  |
|  | Days earlier | *r*<sup>a</sup> | *P* | Days earlier | *r*<sup>a</sup> | *P* | Days earlier | *r*<sup>a</sup> | *P* | Days earlier | *r*<sup>a</sup> | *P* |
| **Covid heart** | 12 | 0.979 | <0.001 | 19 | 0.970 | <0.001 | 12 | 0.982 | <0.001 | 19 | 0.977 | <0.001 |
| **Covid pneumonia** | 14 | 0.978 | <0.001 | 19 | 0.958 | <0.001 | 12 | 0.977 | <0.001 | 19 | 0.967 | <0.001 |
| **Covid** | 12 | 0.978 | <0.001 | 19 | 0.963 | <0.001 | 13 | 0.973 | <0.001 | 20 | 0.972 | <0.001 |
| **Cough** | 19 | 0.932 | <0.001 | 20 | 0.923 | <0.001 | 19 | 0.935 | <0.001 | 20 | 0.945 | <0.001 |
| **Coronavirus** | 19 | 0.914 | <0.001 | 23 | 0.905 | <0.001 | 19 | 0.909 | <0.001 | 22 | 0.925 | <0.001 |
| **Pneumonia** | 19 | 0.848 | <0.001 | 22 | 0.854 | <0.001 | 19 | 0.832 | <0.001 | 22 | 0.897 | <0.001 |
| **Covid diabetes** | 18 | 0.821 | <0.001 | 19 | 0.816 | <0.001 | 18 | 0.812 | <0.001 | 19 | 0.801 | <0.001 |
| **High temperature** | 17 | 0.681 | <0.001 | 22 | 0.641 | 0.006 | 16 | 0.667 | <0.001 | 22 | 0.650 | 0.005 |

<sup>a</sup>The Highest correlation coefficients among the correlation coefficients of a given search term by various days of lagging.

**Figure 1.**
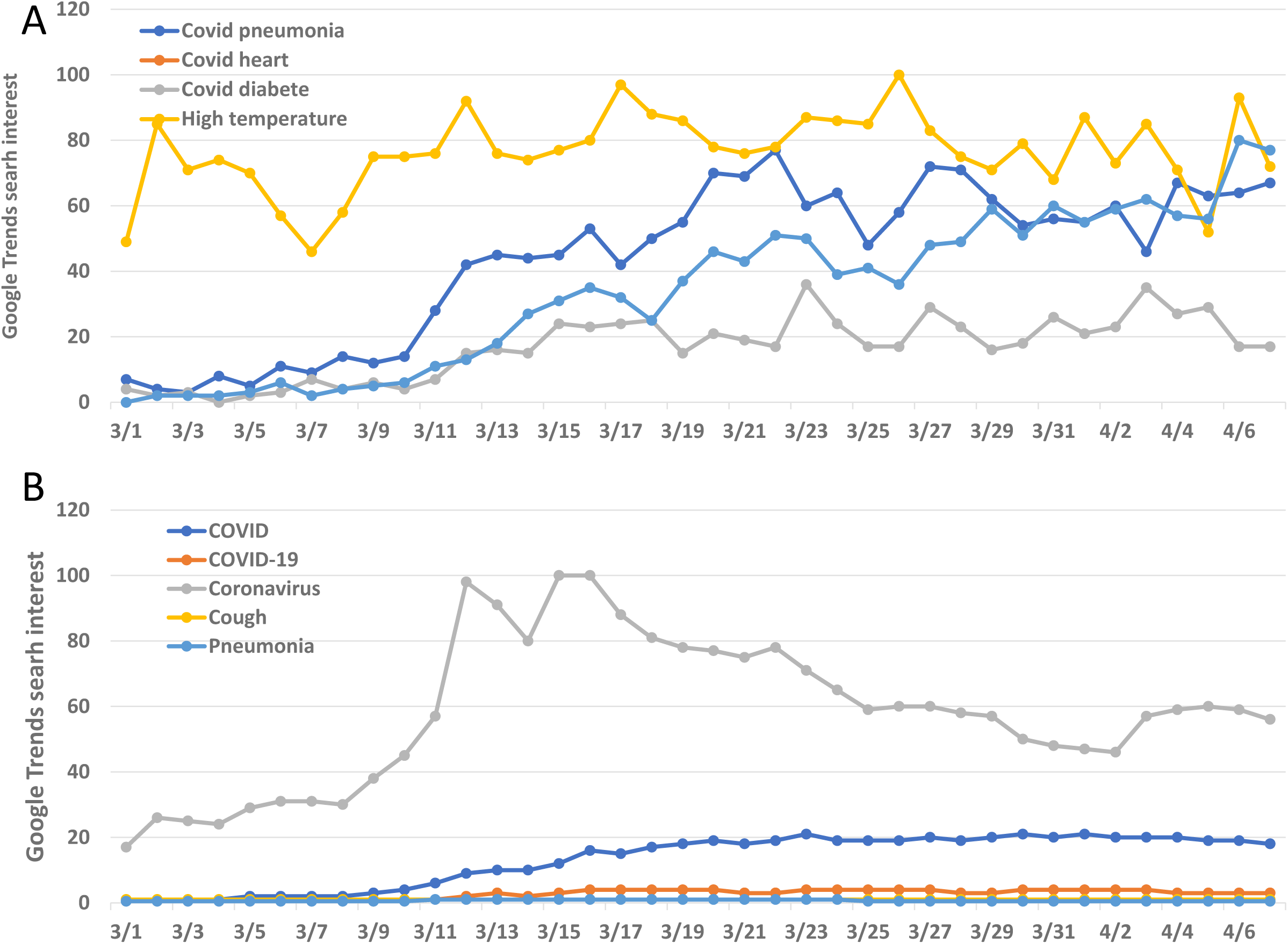
Trends in search interest of COVID-19 related terms. Note: The numbers represented the search interest relative to the term of the highest search interest in the U.S. from March 1 to April 7, 2020.

**Figure 2.**
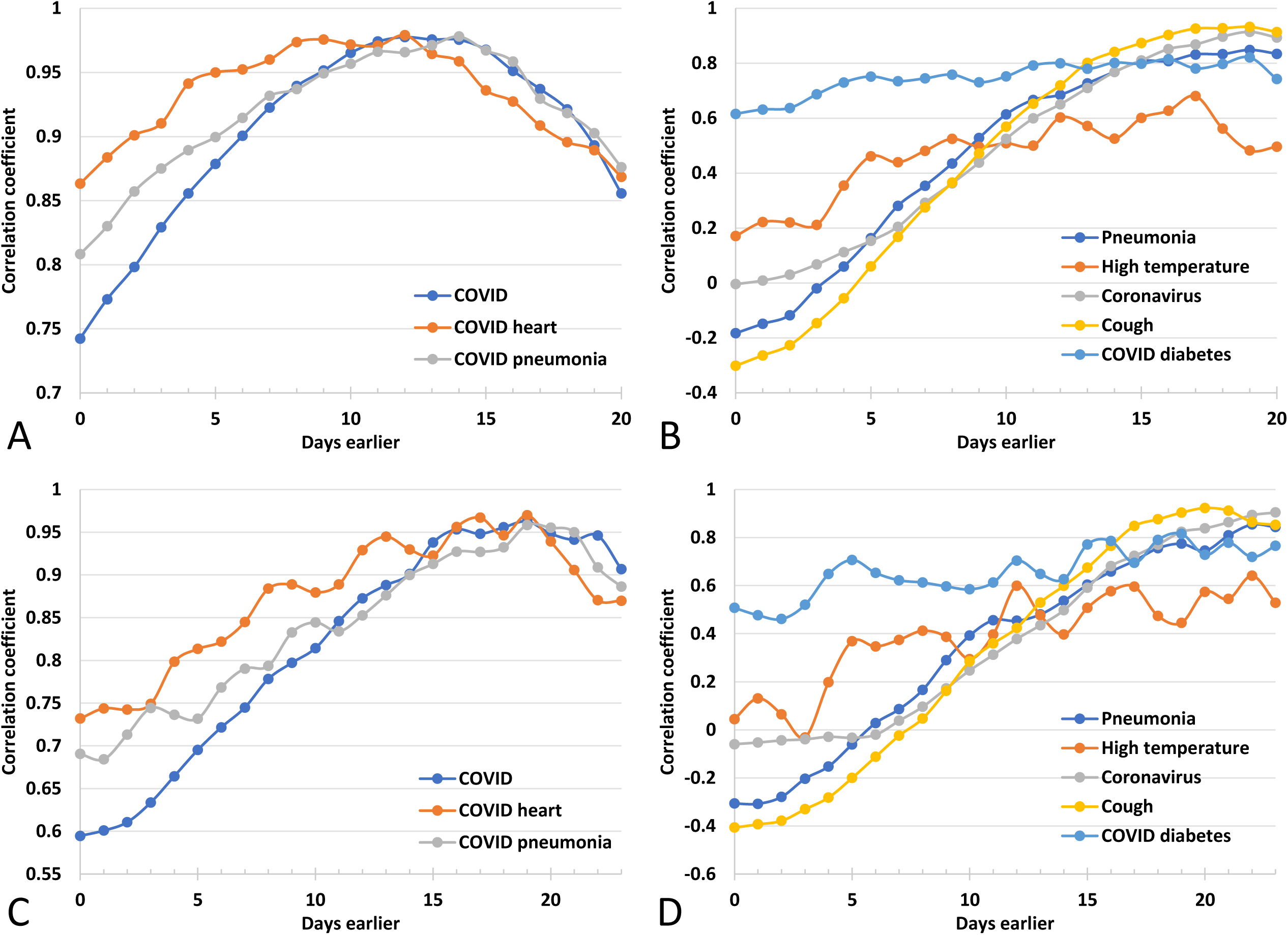
The lag correlations between Google Trends search interest of the terms “COVID,” “COVID heart” and “COVID pneumonia,” and the daily new cases and deaths of COVID-19 in the US, March 1–April 8, 2020. Note: A and C represent the search terms with the highest Pearson’s correlation coefficients for daily incidence and new deaths, respectively. B and D represent the rest of the search terms.

**Figure 3.**
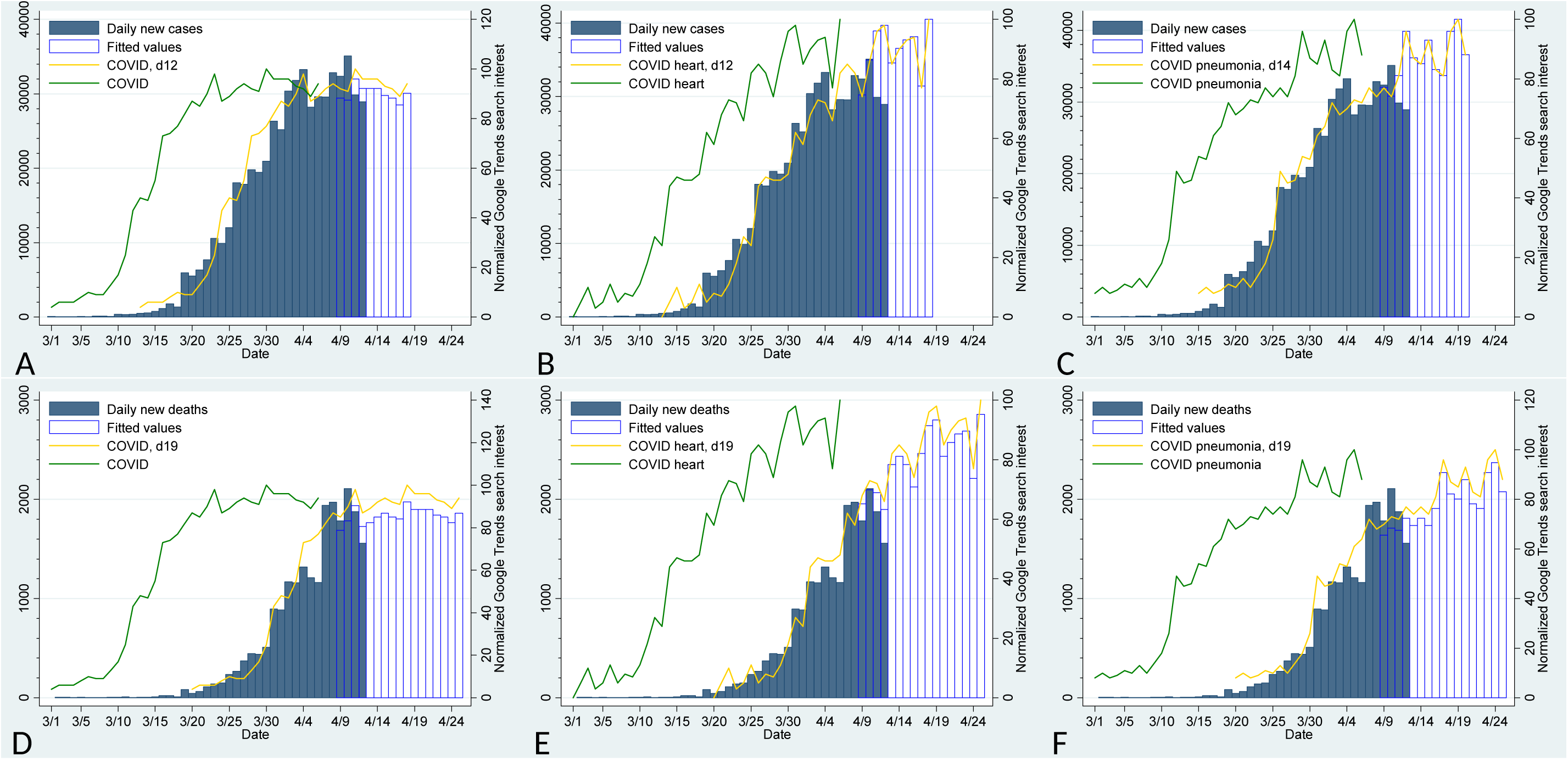
Google Trends search interest and the trends in COVID-19 daily incidence and new deaths in the U.S., March 1 to April 12, 2020. **A-C**. The search interests of COVID, “COVID heart” and COVID pneumonia” in Google Trends were 12 to 13 days lagged from COVID-19 daily new cases/incidence (Pearson’s r= 0.977, 0.982 and 0.973, respectively, *P*<0.001 for all). **D-F**: The search interests of COVID, “COVID heart” and COVID pneumonia” in Google Trends were 19 to 20 days lagged from COVID-19 daily new deaths (Pearson’s r= 0.967, 0.977 and 0.972, respectively, *P*<0.001 for all). Note, d12, d13, d19 and d20 indicate the trend curves were shifted for 12, 13, 19 and 20 days, respectively, to compensate being lagged. The 4-day follow-up with prospectively collected data show Pearson’s r’s were −0.641, − 0.811 and −0.833 for predicting new cases with COVID heart, COVID and COVID pneumonia, respectively, and 0.365, 0.935 and −0.495 for predicting new deaths, respectively.

## Discussion

This population-based study shows that there were 555,245 new cases, and 22,019 deaths of COVID-19 reported in the U.S. from March 1 to April 12, 2020. It also shows that the search interest of COVID, “COVID pneumonia,” and “COVID heart” were highly correlated with COVDI-19 daily incidence and new deaths, with a delay of 12 days and 19 days, respectively.

This study to our knowledge for the first time provided the evidence that search interest pertinent to COVID-19 is highly correlated with the trends in COVID-19 daily incidence and new death in the U.S. The prediction based on 3 search interest items were moderately to greatly accurate in a short-term follow-up study, while additional studies are needed to validate our findings and improve the prediction accuracy. The findings of our study would enable us to better predict new cases and death in the U.S. for the next 12 days, and will greatly help prevent and prepare upcoming pandemic and burdens of COVDI-19 in the future.

The 12 days of lagging time in U.S. as shown by us was longer than previously reported 9 days in China.^7^ Several reasons may contribute to the difference, but should be subject to additional studies. First the testing rate in the United States is much lower than in China. Therefore, many cases may not be tested, and the daily incidence may be underestimated. Second, there was a significant delay in testing for COVID-19 in the U.S. which subsequently led to longer lagging time between the trends of search interest and daily incidence. Third, the biological and socioeconomical differences between the U.S. and Chinese patients may also contribute it to the difference. Finally, the prevalent virus subtypes in U.S. may also be different from that in China and resulted in different lead time.^14^

This study provides several lines of valuable evidence. First, COVID-19 daily new deaths in the U.S. are poorly understood, but are here described and predicted using a semiparametric model. Second, we extensively examined the 9 COVID-19 related search terms, which are more than the 2 used in a previous study.^7^ Our data also suggest that pneumonia and heart problems were highly relevant to the daily incidence and deaths in the U.S. It may be explained by the frequent pneumonia and cardiac injury seen in COVID-19 patietns.^15,16^ Third, the lagging time in our study was longer than the previously reported in China (12 days vs 9 days). However, the 12 and 19 days of lagging time also afforded us the possibility of predicting a longer period of future trends. Fourth, the comparison of predicted values and prospectively collected data will significantly reduce the recall and selection biases. We will continue updating the models’ accuracies as more data become available (see https://github.com/thezhanglab/COVID-US-google). Finally, we rigorously examined the correlation of search interest with the COVID-19 daily incidence and deaths, and show greater correlations (Pearson’s r>0.97) than reported in Chinese data.^7^

This study is limited by the retrospective nature of the modeling part, and may have some related biases. Moreover, due to the relatively low testing rate in the U.S., the daily incidence and deaths may be underestimated. Our sensitivity study using the 1.3 acres data, however, confirms a similar correlation off and search interest with COVID-19 daily incidence and deaths in the U.S.

## The hypothesis and future direction

This population-based, retrospective study show that search terms related to COVID-19 are highly correlated with the trends in daily incidence and new deaths of COVID-19 in the U.S. The prediction-models based on the search interest trend reached moderate to good accuracies. Additional studies are warranted to validate and improve these models.

## Data Availability

Data will be deposited at Github.

https://github.com/thezhanglab/COVID-US-google

## Acknowledgement

The works were in part supported by NIH (R01DK119198 to N.G. and L.Z.). The data will be regularly updated at https://github.com/thezhanglab/COVID-US-google, as new prospectively collected incidence data become available.

## Author Contributions

Dr Zhang had full access to all of the data in the study and takes responsibility for the integrity of the data and the accuracy of the data analysis. They are both senior authors. Drs Yang and Xu both contributed equally and should be considered co-first authors.

Concept and design: Zhang, Gao.

Acquisition, analysis, or interpretation of data: All authors.

Drafting of the manuscript: Yuan and Xu.

Critical revision of the manuscript for important intellectual content: All authors.

Statistical analysis: Yuan, Zhang. Supervision: Zhang.

## Notes

### Competing Interest Statement

The authors have declared no competing interest.

